# A preliminary model to describe the transmission dynamics of Covid-19 between two neighboring cities or countries

**DOI:** 10.1101/2020.07.18.20156695

**Authors:** Raúl Isea

## Abstract

We present a mathematical model that would allow one to describe the transmission dynamics of Covid-19 between two neighboring cities or countries. This model is analyzed both analytical and numerically. It is a preliminary model because it assumes that the migration rate and the mortality rate are constant over time. Despite these simplifications, only two of the four equilibrium conditions were deduced from the system of equations proposed in this paper. Finally, we show an example the transmission dynamics between Portugal and Spain according to the cases registered before June 3, 2020.

## Introduction

Currently the world is suffering from a pandemic caused by a new coronavirus (2019-nCoV) that was detected for the first time in a Chinese market in Wuhan in December, 2019. Up to June 3, more than 7 million infections have been registered worldwide and more than 400,000 have deceased according to data registered at Johns Hopkins University.

To date, a large number of mathematical models have been published to understand the transmission dynamics of Covid-19 in a given community, state, or a country [see for example 1-4]. However, these studies do not consider the influence that the migration effect between neighboring countries can exert, and thus be able to design public policies that best fit the cases registered across the borders.

This type of model is not new in the scientific literature, although there are few cases that analyze real data and have not been used to describe the transmission of Covid-19. In fact, one of the pioneering works was that of Ahmed *et al* [5] who present one of the first models with displacement between two different regions, and later generalized between different n patches by Sattenspiel *et al* [6]. In fact, Wallen and Zhao present various variants based on a SEIR and SEIRS type compartmental model [7,8]. However, it was Chen *et al* [9] who proposed a transmission model where people can become infected during travel.

Unfortunately, the countries do not separate the cases of contagion by local (or community) Covid-19 and those caused by the effect of migration (or imported). This is a limitation that it is necessary to indicate in the future.

Therefore, this work presents a preliminary mathematical model that could explain the transmission dynamics of Covid-19 that moves between neighboring countries as will be explained in the next section.

## Mathematical model

The model is of the compartmental type capable of describing the transmission dynamics between two neighboring cities or countries. The cities are denoted with the subscripts 1 and 2, and we propone the following system of equations:

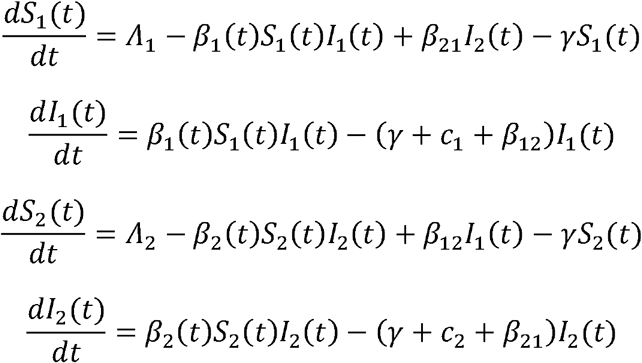

where S_1_(t) and S_2_(t) represent the populations that are susceptible to contracting the virus, while the infected populations are indicated with I_1_(t) and I_2_(t).

This model is based on the following assumptions:

- There is no distinction between local and imported cases in each of the countries since there is no complete record in the databases available (ie., Johns Hopkins University).
- The number of cases of contagion imported from one country to another is considered to be a constant over time. It’s a simplification and therefore the fraction of the population that travels from country 1 to country 2 is represented as *β*_12_, while from 2 to 1 is represented as *β*_21_
- Each country has its own demographic rate and it is equal to *Λ*_1_ and *Λ*_2_.
- Both countries have the same natural mortality rate (=0,00004).
- The mortality rates caused by the Covid-19 are represented as c_1_ and c_2_, and are assumed to be constant over time.
- The contagion rate in each country will be represented with the variables β_1_(t) and β_2_(t), as explained in a recent work published by Isea [4]. These variables are equal to 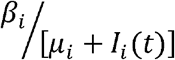 where the subscript represents each of the countries (i = 1,2). The β_i_ and μ_i_ values are determined from a least squares adjustment of the cases of contagion registered in the John Hopkins University database.

In the next section, the system of equations proposed in the work will be analytically investigated.

### Analytical resolution

The methodology required to solve the system of equations proposed in this work has been described on multiple occasions throughout various scientific works [10-14], and for this reason, only we shows the results.

Four steady-state solutions are obtained when the system of equations is solved (denoted as PC^i^, i = 1 until 4) and they are:

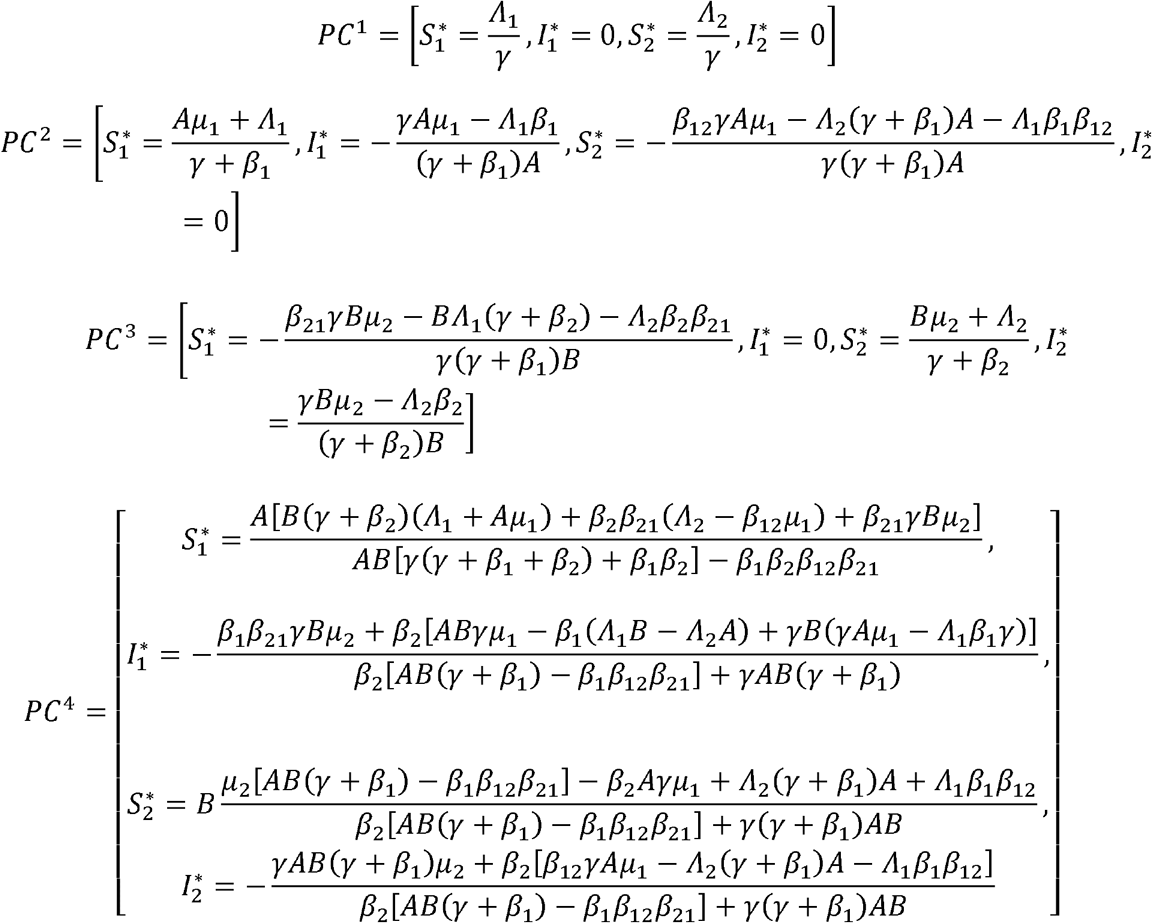

where were defined 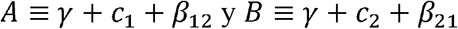.

The next step is to determine if these values (ie., PC^i^, i=1..4) are stable over time, and for this, the Jacobian of the system of equations must be calculated, and evaluated in each of the four solutions indicated above.

The Jacobian obtained from the system of equations (abbreviated as J) is equal to:

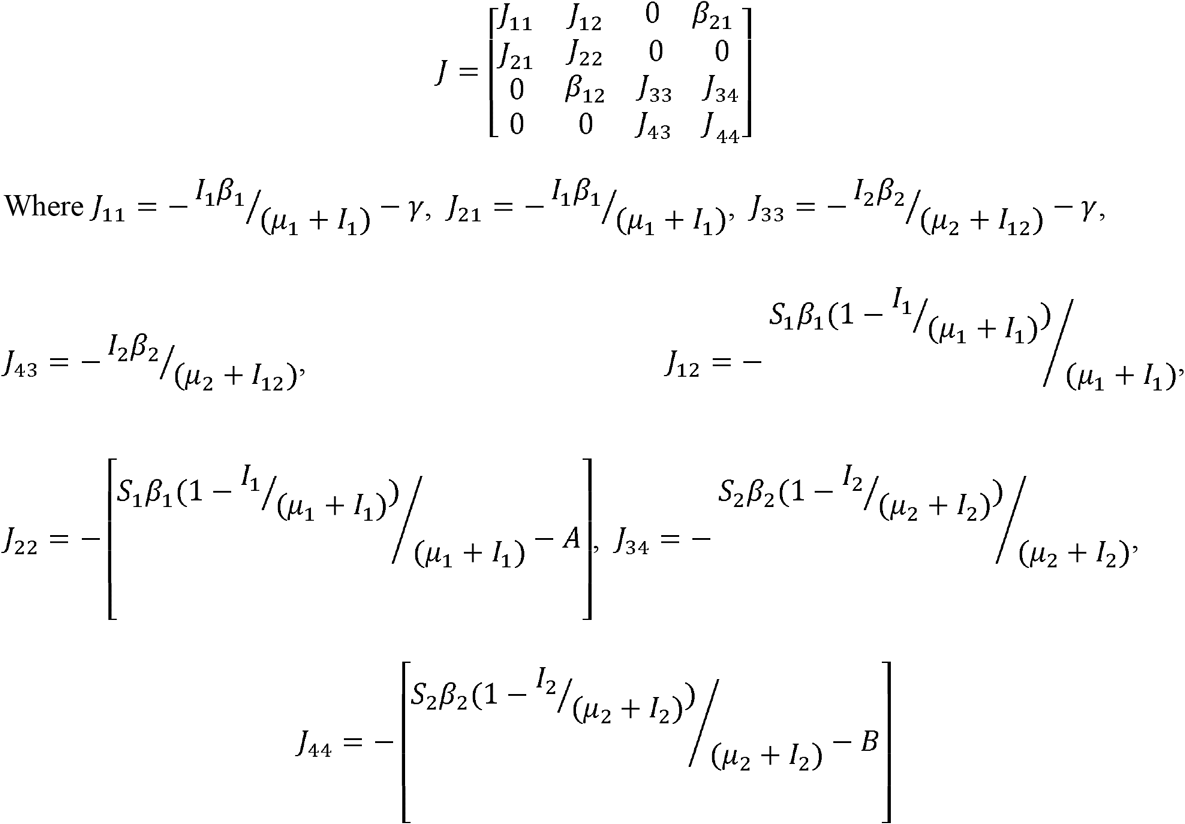

To determine if each solution is stable over time, we must evaluate Jacobian for each of the Critical Points, and later determine the eigenvalues. However, it was only possible to calculate the eigenvalues of the first two critical points (PC^1^ and PC^2^) given that the last two have generated very complex analytical expressions which are difficult to analyze in this paper.

Two eigenvalues that were determined for the first critical point (PC^1^) are:

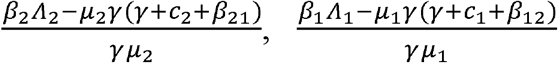

The eigenvalues of the second critical point (PC^2^) are:

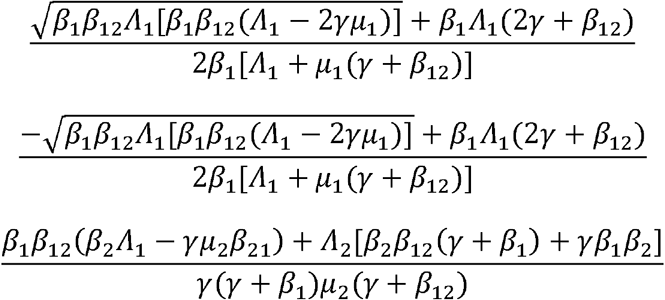

From these values, we can deduce, for example, that *Λ*_1_ > 2*γ μ*_1_, so that the solutions are real. and the next step we shows an example between Portugal and Spain.

### Numerical solution

We analyzed the number of infected cases between Portugal and Spain, denoted as 1 and 2, respectively. To do that, it is necessary to fit using the least squares method for each of the parameters of the system of equations based on the data recorded in the Johns Hopkins University up to June 3, 2020. We have preferred to adjust the data country by country because we don’t have data separating the local and imported cases between these countries.

The figure 1 shows the results obtained in Portugal where the data are represents with circle and the least squared adjusted with a line. The normalized values obtained in Portugal are *Λ*_1_ = 5.540,: *β*_1_ = 0.103, *μ*_1_ = 0.162; while that the results obtained for Spain are *Λ*_2_ = 0.026 *β*_2_ = 0.027, *μ*_1_ = 0.044.

**Fig 1.**
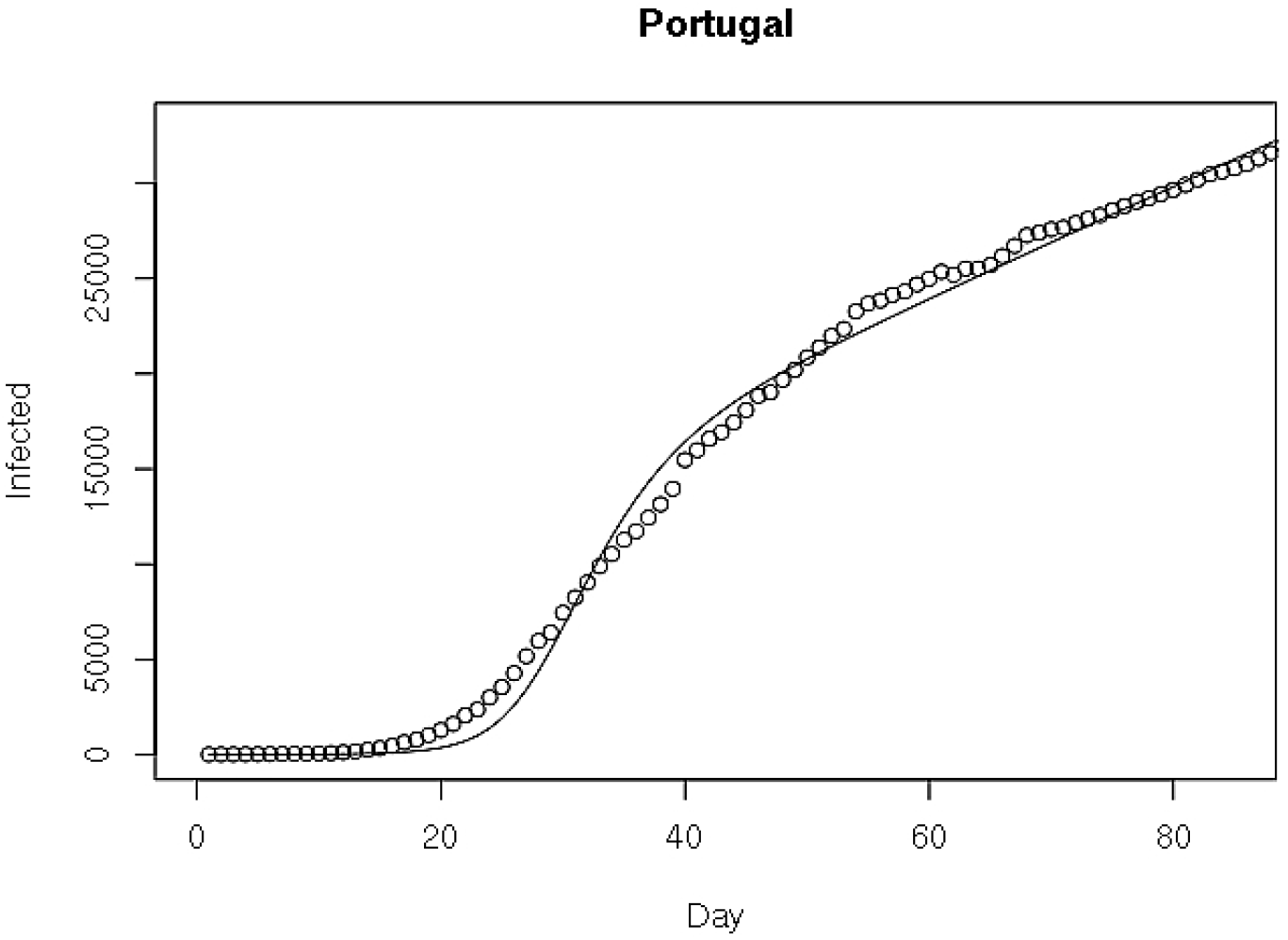
Cases of Covid-19 in Portugal (circle) and the results obtained by least square adjusted (line). This result was obtained with a Python program.

The figure 2 shows the vector field diagrams of the infected cases in each of the countries (I_1_ versus I_2_) that were evaluated with the parameters obtained in the previous step. As can be seen in Figure 2(a), the transmission dynamics of Covid-19 between the countries when there are no imported cases (β_12_ = β_21_ = 0). Figure fig 2(b) considers that there is a migration of people from Portugal to Spain (B_12_ = 50, B_21_ = 0. These values were randomly selected. Finally, Figure 2(c) presents an extreme example where a high migration of people from Portugal to Spain is assumed highlighting the exponential growth of cases in Spain.

**Figure 2(a).**
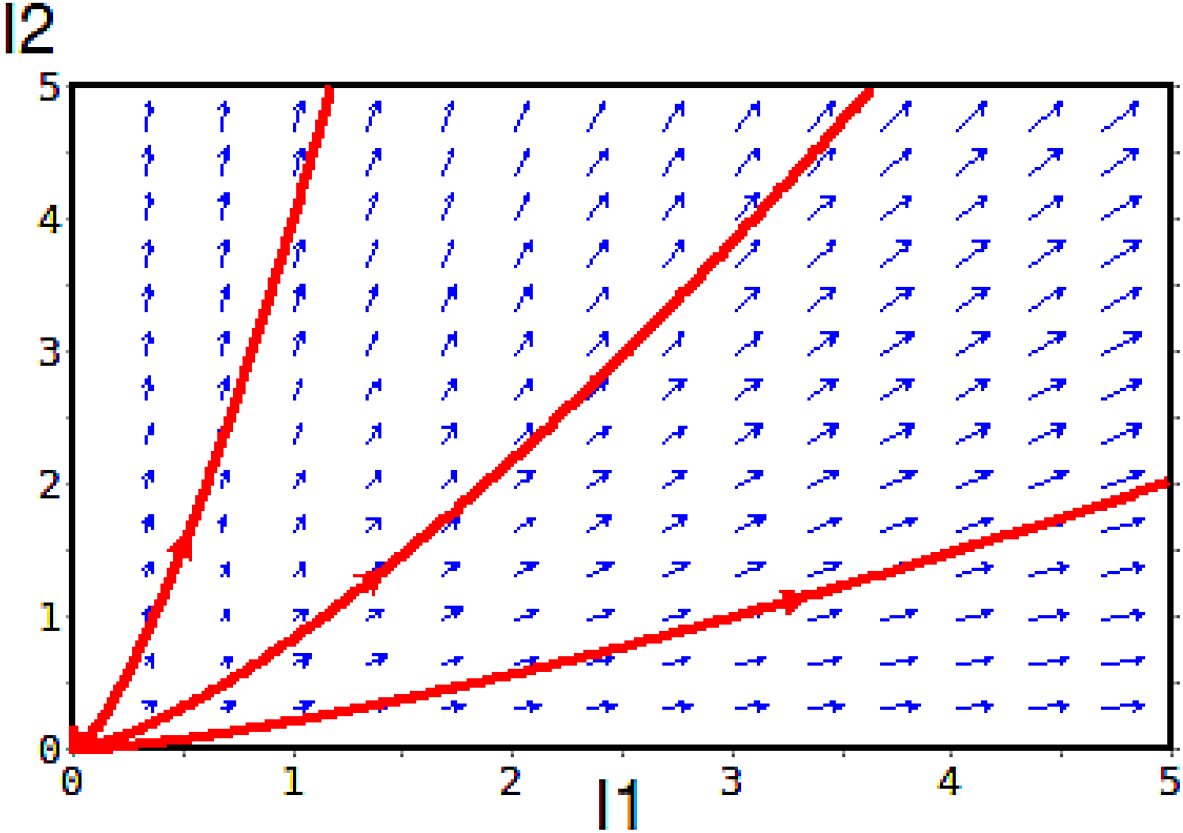
Vector field diagram of Covid-19 infected cases between Portugal and Spain (denoted as 1 and 2, respectively). In this example, we considered an initial susceptible population S_1_ = 100 and S_2_ = 140.

**Fig 2(b).**
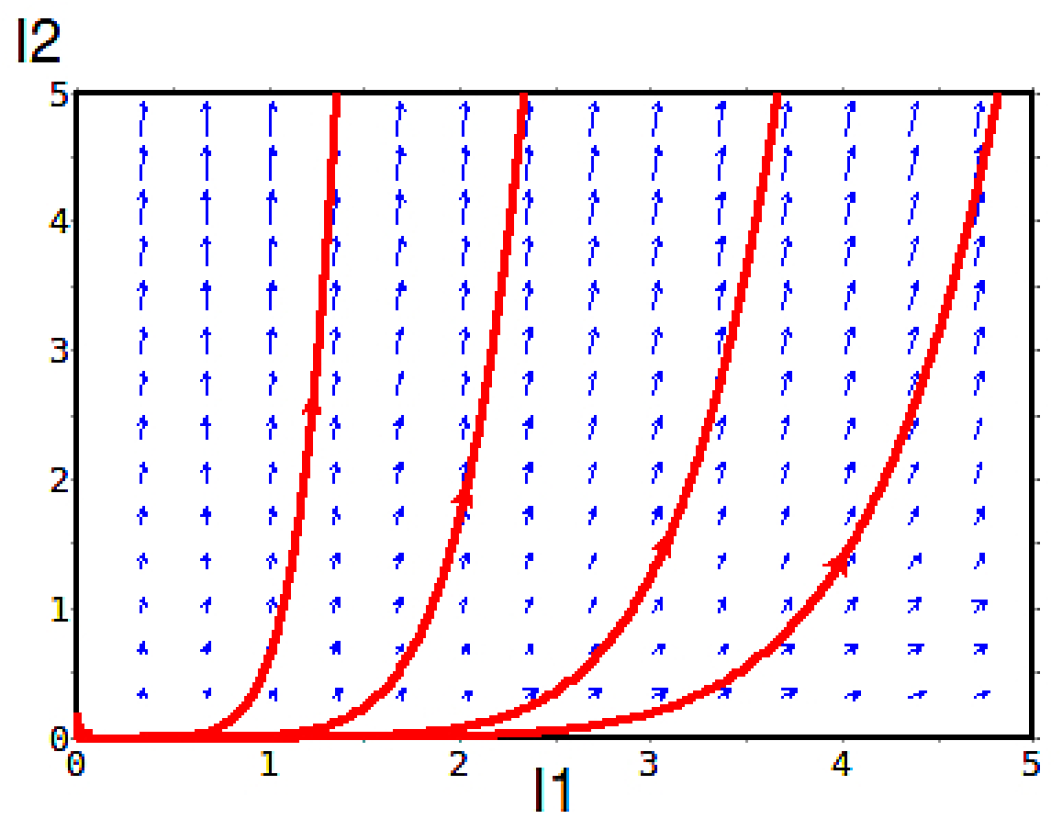
Vector field diagram where there is only a migration of imported cases from Portugal to Spain (B_12_ = 50, B_21_ = 0) where it is seen how the cases of contagion in Spain increase significantly.

**Fig 2(c).**
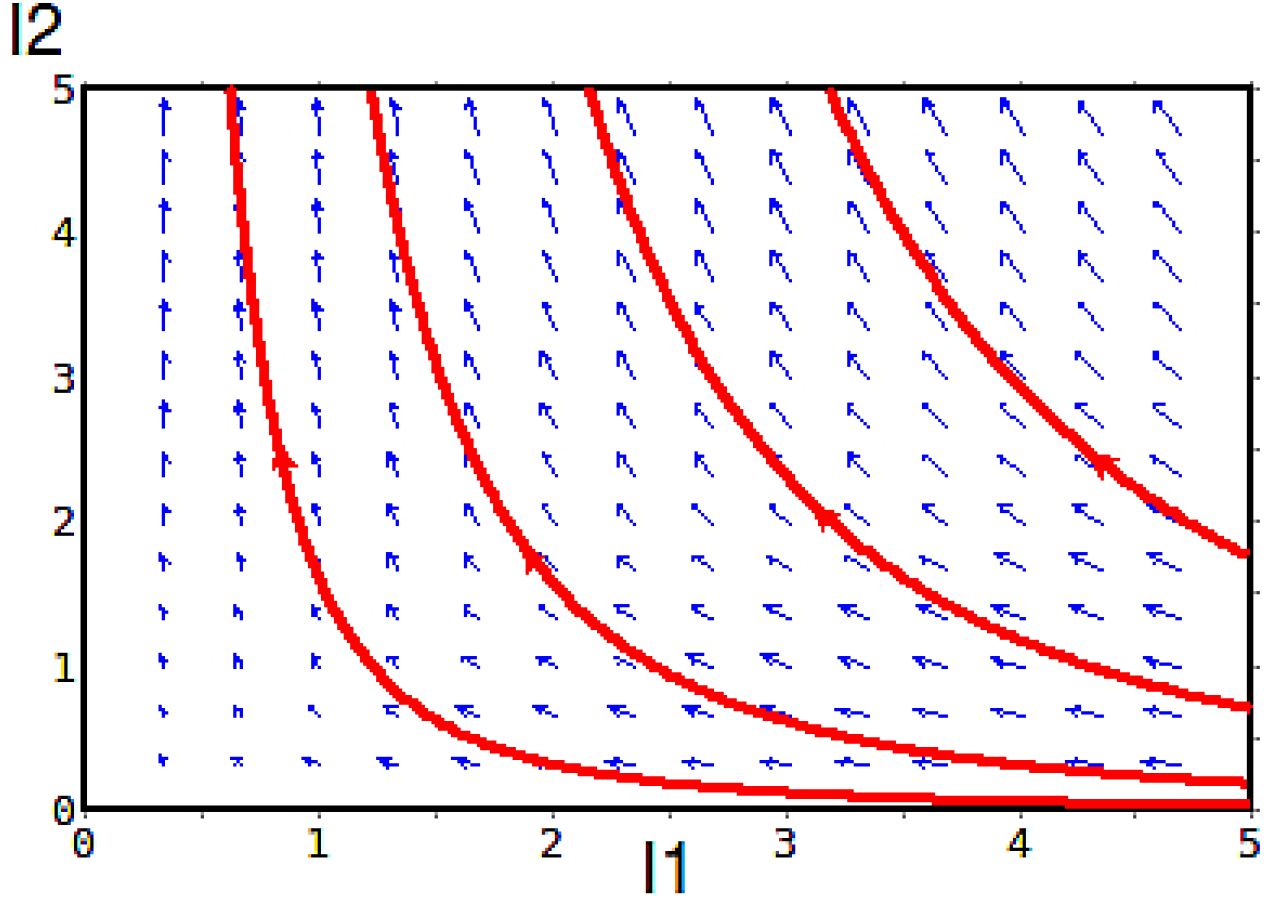
It is an extreme example where it is only considered that there could be a high migration of cases of infections that leave Portugal to Spain (B_12_ = 100, B_21_ = 0), and we verify that Portugal presents few cases in comparison to Spain.

## Conclusions

The present mathematical model proposes a system of differential equations that will allow us to understand how the transmission dynamics of cases by Covid-19 varies between two neighboring countries. This system was partially solved analytically due to the complexity of the solutions found in the work. Finally, an example of how the number of cases of Covid-19 infection between Portugal and Spain can vary is presented.

It is necessary that the countries must separate the local cases from the imported ones, and in this way be able to propose more realistic models, and in this way, for example, be able to relax or radicalize the measures at the borders between neighboring countries.

## Data Availability

It is a mathematical model. All data in Jonhs Hopkins University about Covid-19

## Acknowledgment

I’d like to acknowledgment to Karl E. Longreen for your comments in this manuscript.

